# Characterization of an extreme phenotype of schizophrenia among women with homelessness

**DOI:** 10.1101/2023.07.29.23293378

**Authors:** Jayakumar Menon, Suvarna Jyothi Kantipudi, Aruna Mani, Rajiv Radhakrishnan

## Abstract

**Background:** Studies of schizophrenia and homelessness are often confounded by comorbid substance use. Women with schizophrenia and homelessness in India have very low rates of substance use and provide a unique opportunity to disentangle the effects of illness from that of substance use. We examined the clinical characteristics of women with schizophrenia and homelessness and compared it to an age-matched group of women with schizophrenia living with their family.

**Methods:** 36 women with schizophrenia and homelessness, and 32 women with schizophrenia who were illness living with family were evaluated for psychopathology using Scale for Assessment of Positive Symptoms (SAPS)/ Scale for assessment of negative symptoms (SANS) scales, cognitive difficulties using Montreal Cognitive Assessment (MOCA)/Rowland Universal Dementia Scale (RUDAS), and Frontal Assessment Battery(FAB), disability using World Health Organization - Disability assessment Scale (WHO-DAS) and psychosocial factors using a semi-structured proforma. The groups were compared using t-tests and chi-square for continuous and categorical variables respectively.

**Results:** Women with schizophrenia and homelessness were found to have significantly higher scores on measures of psychopathology, significantly lower cognitive functioning, and much higher disability, and were also on higher doses of antipsychotics. The mean scores on measures of psychopathology, cognition and disability for women with schizophrenia and homelessness differed by 2-3 standard deviations with the mean for women living with family (i.e. z scores) suggesting that they represented an extreme phenotype. Rates of past employment were higher among women with schizophrenia and homelessness. Hence these differences were not accounted for by premorbid functioning.

**Conclusions:** The study raises the possibility of an extreme phenotype of schizophrenia with severe and persistent psychopathology non-responsive to dopamine blocking drugs, cognitive impairment, and disability, which needs further exploration.

## INTRODUCTION

Schizophrenia is a major psychiatric disorder known to impact multiple domains of an individual’s life and lead to poor outcomes including homelessness and disability. ^(1,2,3)^. Homelessness and schizophrenia are thought to have a bi-directional relationship ^(4)^. Wandering schizophrenia patients are also known to have more severe psychopathology, probably owing to poor treatment adherence ^(5,6)^. In male patients, higher severity of positive symptoms and to a lesser extent substance abuse and medication non-adherence is associated with homelessness^(6,7)^.

India has widely different mental health service delivery across its states. The government supported national programs, National Mental Health Program (NMHP) and District Mental Health Program (DMHP) primarily addresses the acute treatment gap^(8)^. Metropolitan cities have the greatest number of psychiatrists and mental health establishments. There is hardly any community mental health service to support patients once discharged from acute care, and family still takes up the entire burden of care in this society compared to higher-income countries. Increasing urban migration, nuclear family system in the cities, loss of traditional community systems in rural areas, rising costs of living without commensurate increase in income, and lack of adequate social support like disability pension, unemployment benefits and social housing options are leading to a gradual but steady decline in care for the serious mentally ill^(9,10)^.

Studies of schizophrenia with coexisting homelessness are often confounded by the high rates of substance use in this population Women with schizophrenia in low and middle-income countries including India, although thought to have better outcomes, are more prone for poor family support and homelessness ^(11).^ Interestingly, this population in India has very low rates of substance use. This provides a unique opportunity to disentangle the effects of illness from that of comorbid substance use. However, women with schizophrenia and homelessness have not been studied adequately in India^(11,12)^. Not much is known about the severity of illness, response to treatment, cognitive deficits, and functional impairment in this group, which by itself could be reasons for the poor outcome. In this context, the current study aimed to compare psychopathology, socio-occupational functioning, disability, and cognitive functioning among women with schizophrenia and homelessness and age, gender matched controls with schizophrenia who were living with family.

## METHODS

### Study Design

This was as a cross-sectional observational study of two groups of female patients diagnosed with schizophrenia (those with homelessness and those living with family), comparing symptom severity, cognitive functioning, and disability. Principles of observational studies as per STROBE guidelines were used to decide on the design and description of data. The study was approved by the Institutional Ethics Committee, Sri Ramachandra Institute of Higher Education and Research (SRIHER), Chennai (Ref:IEC/NI/20/OCT/76/99).

### Settings

Patients were selected from 2 different settings; those living with family support in their homes and those living in a long stay home for mentally ill women with homelessness, who were rescued from the streets. The home is being run by a non-government organization (NGO) involved in rescue, rehabilitation and reintegration of mentally ill individuals, Anbagam-TERDOD. The control group, those living with family, were selected from the tertiary hospital’s psychiatry out-patient clinic (SRIHER, Chennai). The socio-demographic variables were collected using a semi-structured questionnaire, and further details were also obtained from the patient records. The patients were recruited from January 2020 to June 2022.

### Participants

Patients with family support were selected from the outpatient clinic of a tertiary care hospital. The hospital caters to the population in the western parts of the city, and mostly encompasses patients from lower- and middle-income families. Convenience sampling was used, and consecutive patients were selected after review by the consultant psychiatrist, once they fulfilled the inclusion criteria. Senior consultant psychiatrist (SJK) confirmed the diagnosis through detailed interviews and verification of clinical records.

Women with schizophrenia and homelessness were selected from the not-for-profit organization run rehabilitation home (Anbagam-TERDOD) situated in the same city. All the female patients were rescued from the streets, in a wandering, unwell state by the Police and civic authorities and subsequently placed in the process of treatment and rehabilitation. The patients were evaluated by a senior consultant psychiatrist (JM), carried a diagnosis of schizophrenia and had been on antipsychotic treatment for at least 6 months as per established treatment guidelines. The patients included were predominantly Tamil speaking and a few were Hindi speaking. The investigators were fluent in both these languages as well as English.

### Variables

The following were the variables of interest: 1) psychosis severity as assessed by Scale for Assessment of Positive Symptoms (SAPS) and Scale for Assessment of Negative Symptoms (SANS). SAPS/SANS is a well validated scale used universally for evaluation of psychopathology in schizophrenia and related disorders^(13)^; 2) Cognitive function assessed by Montreal Cognitive Assessment (MOCA) for literate patients / Rowland Universal Dementia Assessment Scale (RUDAS) for illiterate patients and Frontal Assessment Battery (FAB)^(14,15,16)^. MOCA has validated versions in Tamil and Hindi and hence it was used for assessment of the patients. RUDAS is language and culture independent scale for screening of cognitive functions and can be administered in various clinical settings. Both scales have been previously used in patients with schizophrenia and noted to be useful in assessing cognitive function in long standing psychosis. ^(17)^. 3) Disability due to the illness was assessed by WHO-DAS ^(18,19)^.

### Data Sources/Measurement

For the women with schizophrenia and homelessness, living in the long stay shelter, senior consultant psychiatrist (JM) evaluated the patients using PANSS interview schedule and subsequently scored the SAPS/SANS. A trained mental health nurse fluent in the language and with expertise in administering screening tools for cognition and disability completed the assessments with MOCA, FAB and WHO-DAS. Randomly, some of the patients were independently assessed by the senior consultant psychiatrist (JM) in order to ensure consistency in assessment and scoring of scales.

Women with schizophrenia living with family were evaluated by senior consultant psychiatrist (SJK) and verified the diagnosis and treatment. A trained clinical psychologist conducted the structured clinical interview using PANSS interview schedule, and additional questions were added to cover the entire SAPS/SANS scoring. The disability assessment was performed in collaboration with a family member. The cognitive assessments were performed on a follow-up visit (within 2 weeks).

### Bias

The two raters remained blind to the assessment of the other for the entire duration of the study. There was no communication between the two teams about results nor trends until the completion of assessment of the entire patient population in both groups. Even though selection bias was unavoidable, consecutive sampling of patients who fulfilled intake criteria helped in mitigating this.

### Study size

In view of the limited numbers of patients presenting with homelessness and unwell status in the NGO directly, or through transfer, compared to those with supports, it was impossible to gather large samples of the first group. There were no previous studies from India which looked at these variables in detail and hence a sample size was not arrived at a-priori. For statistical evaluation using t-tests, 25 is considered as the minimum number of participants. We were able to complete evaluation of 36 subjects during the study period.

### Statistical methods

IBM SPSS (Statistical Package for Social Sciences) Version 27 was used to do the statistical analysis. The sociodemographic, treatment, illness, cognitive scores, and disability scores were presented in frequencies and mean ± standard deviation. The Chi-square and Fischer exact test were used to assess the significance of the association between categorical variables. The significance of the association between continuous variables was assessed using the independent sample t-test. P value of < 0.05 was taken as a statistically significant difference between the two groups.

## RESULTS

### Participants

A total of 60 consecutive women with schizophrenia living with family support were screened for eligibility, 36 women were found eligible and 32 consented to the study.

Of the 60 women with schizophrenia and homelessness who were screened for eligibility, 45 women were found to be eligible and 36 were recruited to the study following informed consent. 10 patients who lacked capacity because of the severity of the illness, nominated representative gave permission for the assessment, once they were found eligible to participate. The age and illness duration of those who were recruited and those not recruited were not significantly different.

### Descriptive data

The study sample included 68 women with schizophrenia, 32 living with family support, and 36 with homelessness (living in long-stay shelters). The mean age of the women living with support and those with homelessness was 44.5 years (SD 9.5) and 41.39 years (SD 7.9) respectively.

**Table 1** shows the sociodemographic and treatment characteristics of the sample.

**Table 1.**
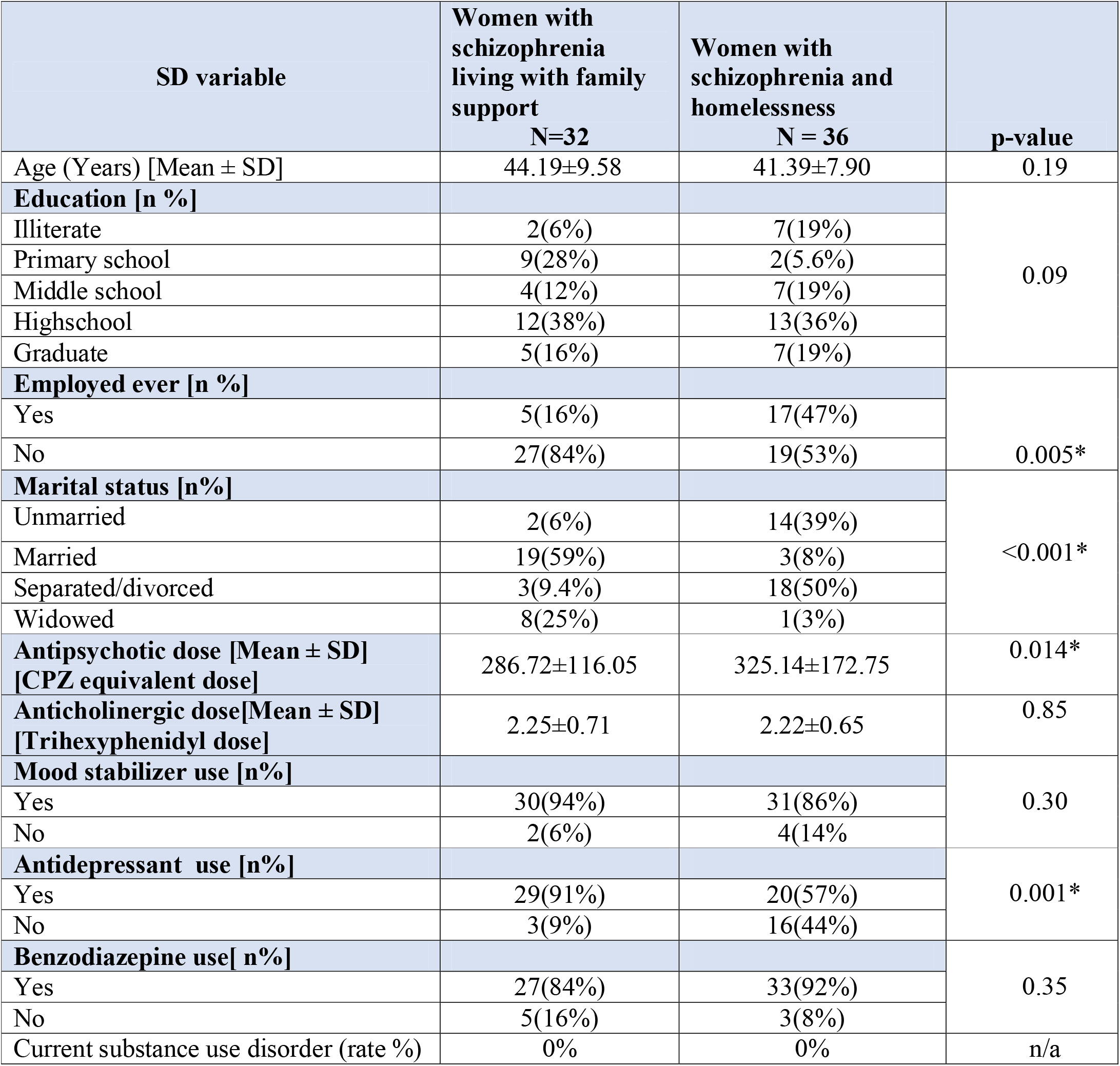
Socio-Demographic and treatment details of the study participants.

#### Outcome data

**Figure 1.**
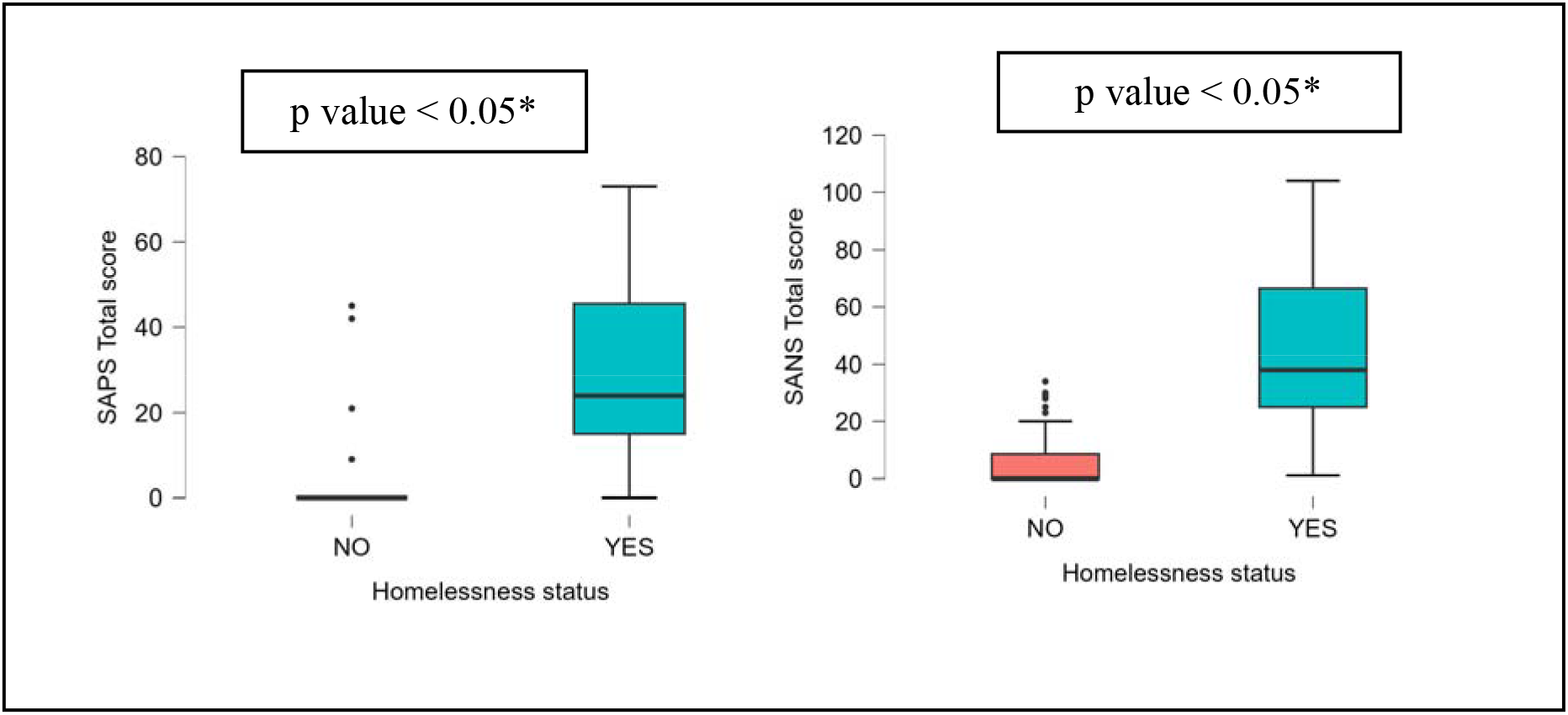
depicts the boxplots of SAPS and SANS total scores by homelessness status

**Figure 2.**
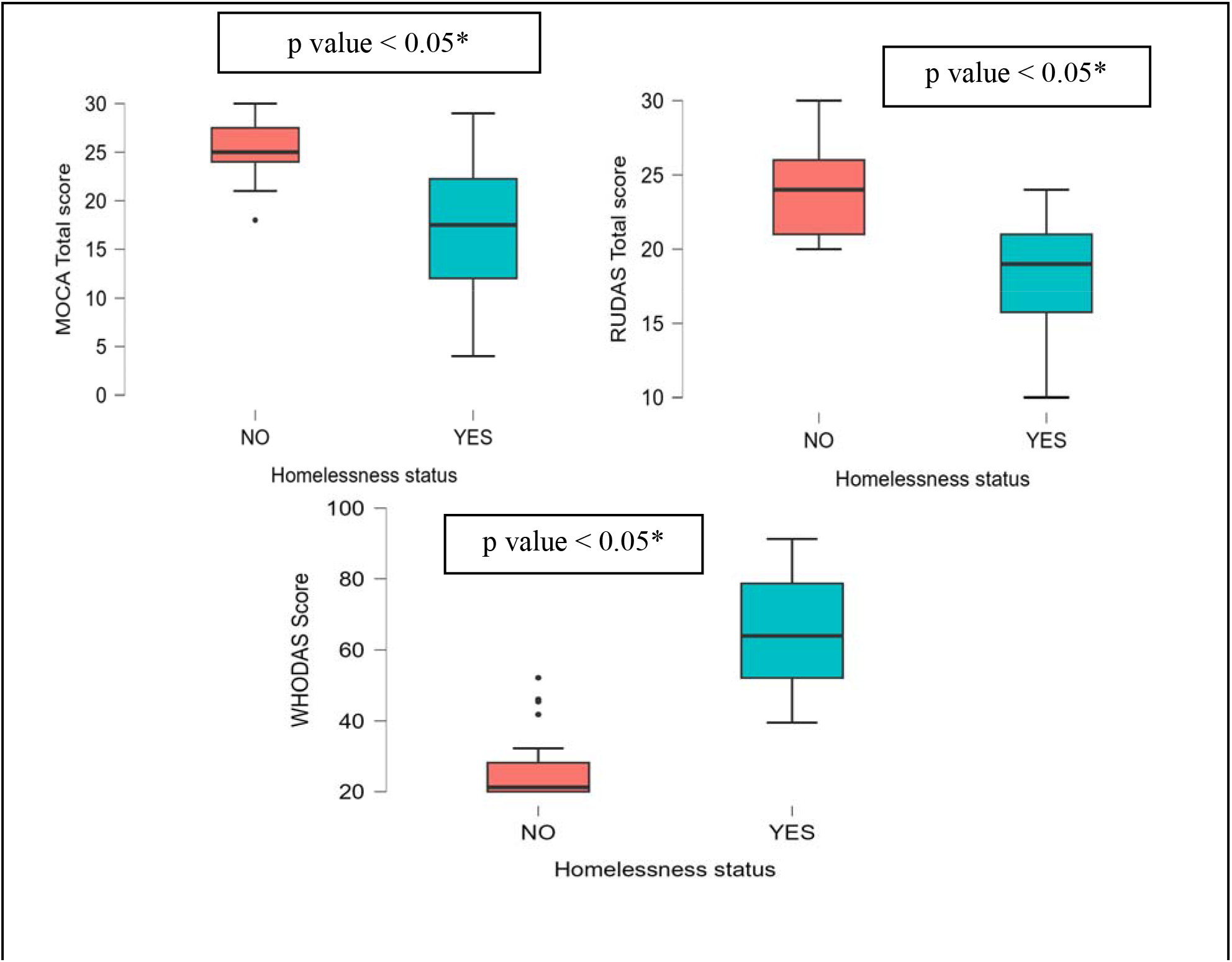
depicts the boxplots of cognitive scores and disability scores by homelessness status.

**Table: 2.** Illness, cognition and disability score details of the study participants.

|  | **Women with schizophrenia living with family support** | **Women with schizophrenia and homelessness** | **p-value** | **Effect size (Cohens d) 95% CI s** | **Z score** |
| --- | --- | --- | --- | --- | --- |
| **SAPS total score**<br>[Mean ± SD] | 3.66±11.19<br>**N=32** | 29.92±19.89<br>**N = 36** | <0.001* | 1.60<br>[1.05 to 2.15] | 2.1 |
| **SANS total score**<br>[Mean ± SD] | 6.66±11.43<br>**N=32** | 46.11±26.92<br>**N = 36** | <0.001* | 1.87<br>[1.29 to 2.44] | 3.5 |
| **MOCA Total score**<br>[Mean ± SD] | 23.32±3.39<br>**N=19** | 16.79±7.01<br>**N=28** | <0.001* | 1.46<br>[2.11 to 0.80] | -1.9 |
| **RUDAS Total score**<br>[Mean ± SD] | 24.31±3.33<br>**N=13** | 17.88±4.76<br>**N=8** | 0.002* | 1.64<br>[2.65 to 0.60] | -1.9 |
| **FAB Total score**<br>[Mean ± SD] | 15.38±1.56<br>**N=32** | 9.64±4.52<br>**N = 36** | <0.001* | 1.66<br>[1.10 to 2.21] | -3.7 |
| **WHODAS percent score**<br>[Mean ± SD] | 23.45±9.37<br>**N=32** | 55.57±12.2<br>**N = 36** | <0.001* | 2.93<br>[2.23 to 3.61] | 3.4 |

## Results

The mean age of the sample was 41.39±7.90 years. The age and education profile of both groups were not significantly different (Table 1). Women with schizophrenia and homelessness had higher rates of lifetime employment compared to women living with family support. Greater proportion of women living with the family were married compared to women with homelessness. The mean antipsychotic dose (chlorpromazine equivalents) among women with schizophrenia and homelessness was significantly higher than those with SZ living with family support. 10 out of 36 patients in the long-stay home were on clozapine, and even in them the response has been less than ideal, with only 20 percent (2 out of 10) showing good clinical and functional improvement on clozapine.

In terms of psychosis severity, women with schizophrenia and homelessness had significantly higher total SAPS scores (Mean±SD = 29.92±19.89) than the women living with family support (Mean±SD= 3.66±11.19, t value 6.56, df 66, Cohen’s d effect-size 1.60, z-score 2.1). They also had significantly higher total SANS scores (Mean±SD = 46.11±26.92) compared to women living with family support (Mean±SD= 6.66±11.43, t value 7.69, df 66, Cohen’s d effect-size 1.87, z-score 3.5).

On cognitive performance, women with schizophrenia and homelessness had significantly lower total MOCA scores (Mean±SD =16 .79±7.01) than the women living with family support (Mean±SD= 23.32±3.39, t value -4.92, df 45, Cohen’s d effect-size 1.46, z-score - 1.9). They also had significantly lower total RUDAS scores (Mean±SD= 17.88±4.76) than the women living with family support (Mean±SD = 24.31±3.33, t value -3.65, df 19, Cohen’s d effect-size -1.64, z-score -1.9), and significantly lower FAB total score (Mean±SD= 9.64±4.52) than the women living with family support (Mean±SD = 15.38±1.56, t value - 6.83, df 66, Cohen’s d effect-size -1.66, z-score -3.7)

In terms of global level of functioning, women with schizophrenia and homelessness had significantly higher WHODAS disability percent scores (Mean±SD= 55.57±12.2) than the women living with family support (Mean±SD= 23.45±9.37, t value -12.06, df 66, Cohen’s d effect-size 2.93, z-score 3.4). The mean domain scores of WHODAS and the mean total disability percent score are higher in women with schizophrenia and homelessness than those of women living with their families and the difference was statistically significant.

The primary result of the study shows significantly greater severity of psychopathology, cognitive impairment, and extent of disability among women with schizophrenia and homelessness. Notably, women with schizophrenia and homelessness had higher rates of lifetime employment compared to women living with family support, suggesting that these differences cannot be explained by premorbid level of functioning. None of the patients had an active substance use disorder. The findings were hence not confounded by comorbid substance use. The control sample in this study was of comparable socio-economic background. The hospital population were from the general psychiatry outpatient clinic which caters to the low and lower-middle socio-economic section of the society. All women with schizophrenia and homelessness belonged to lower socio-economic status. There was no difference in age or educational level of the patient groups. One major demographic difference between the groups was in marital status. Greater proportion of women with schizophrenia and homelessness were never married/separated or divorced from their husbands, which reflects the poorer socio-occupational outcomes of women with psychosis across the world^4^.

## DISCUSSION

To the best of our knowledge, this is the first study from India to examine clinical characteristics of women with schizophrenia and homelessness, compared to a similar age and gender matched group who have family supports. The study found that women with schizophrenia and homelessness, have more severe illness (> 3 standard deviations from the mean of control group measured by SANS, SAPS), greater cognitive deficits (>2-3 standard deviations from the mean of control group) and greater disability ((> 3 standard deviations from the mean of control group). They were also on higher doses of antipsychotic medications (CPZ equivalents). The large degree of deviation (>3 standard deviations) from the control group suggests that women with schizophrenia and homelessness represent an extreme phenotype of the illness. These findings were not confounded by comorbid substance use, unlike prior studies from western countries ^(8)^.

While schizophrenia and homelessness are considered to have a bidirectional relationship ^(6)^, not all patients with schizophrenia experience homelessness. Cognitive impairment has been shown to be an important domain in schizophrenia which impacts outcome ^(20,21,22)^. Study comparing sheltered and unsheltered homeless veterans have shown that overall cognition, visual learning and social cognition were the important factors that associated with longer stay in unsheltered locations ^(23)^. Homelessness could be the outcome of this severe impairment in cognition, which incapacitates the patients in this group to perform even the basic demands of life, leading to decline in socio-occupational functioning.

Our study shows that patients with homelessness have much severe cognitive impairment compared to those living with family, and the difference between the groups were of large effect size. The degree of impairment was to such an extent that patients were unable to participate in detailed cognitive testing using specific cognitive assessment tools used in schizophrenia. Most of the patients in the homeless group could not understand even simple instructions in the Tamil version of the Brief Assessment of Cognition in Schizophrenia (BACS) or NIMHANS battery, two commonly used cognitive assessment tools in schizophrenia. Frontal/executive functions were especially affected as evidenced by the scores both in MOCA and FAB. This is consistent with previous studies on the prevalence and effects of frontal dysfunction in schizophrenia ^(24,25,26,27).^ There was difficulty in carrying out cognitive assessment in the homeless group, possibly owing to the extent of psychopathology and severity of cognitive difficulties in that group. Following extensive discussions with neuropsychologists from National Institute of Mental Health and Neurosciences (NIMHANS) and Schizophrenia Research Foundation (SCARF), who had vast experience in research in patients with schizophrenia, cognitive screening tools MOCA and RUDAS were considered, as they had good construct validity and inter-rater reliability, and validated local language versions were available.

Impairment in cognitive functions was not unexpected, but the extent and severity and the significant difference between the group with a large effect size (1.64-1.66) also supports the notion that this population represents an extreme phenotype of the illness. This is also reminiscent of the concept of hebephrenic schizophrenia (described by Kahlbaum and Ewald Heckers) with unique clinical picture characterized by severe and persistent cognitive impairment, that remains non-responsive to antipsychotic medications ^(28,29).^The clinical outcome remains poor in this subtype^(30)^ and has historically been thought to result in homelessness. It remains to be seen if these patients fare better with treatment initiation earlier in the course of the illness.

The other major finding from the study is the persistence of severe psychopathology (positive and negative symptoms) in the women with homelessness group. Almost all the patients had an adequate trial of 2 different second-generation antipsychotics (SGAs) and many were subsequently started on clozapine. Average duration of each SGA trial was at least 3 months and those who showed minimal or poor clinical response were moved to clozapine. The average dose (chlorpromazine equivalent) of the patients with homelessness were significantly higher. This is in contrast to prescribing practices among patients with homelessness in the US where less than 0.2% veterans with homelessness were treated with clozapine^(31)^.

The higher dose of antipsychotics in this population raises the question of whether this extreme phenotype of patients predominantly suffer from psychosis mediated by non-dopaminergic pathways. Recent research using neuromelanin MRI has brought to light non-dopaminergic subtype of schizophrenia with poor response to SGAs and other dopamine receptor blocking drugs and better response to clozapine ^(32,33,34)^. Follow up studies using such biomarkers in the homeless population to verify this hypothesis maybe a worthwhile endeavor. Similar biomarker studies done in first episode psychosis have attempted to predict those with poor response to both first-generation antipsychotics (FGAs) and SGAs ^(35)^. Clozapine response is better predicted by higher psychosis scores, however it is also imperative to transition to medications with lesser side-effects if there is poor response to clozapine even after 6 months ^(36,37)^. The utility of clozapine in the long-stay home population has been less than ideal, with less than 20 percent treatment-resistant patients responding to clozapine.

The significant impairment in socio-occupational dysfunction in patients with homelessness is another matter that require further evaluation. A combination of social worker led adherence enhancement strategies with long-acting injectable antipsychotics appear to improve outcomes in African-American youth with substance and severe mental illness^(38)^. However in our study, all the patients were under supervised medication and inpatient rehabilitation. The NGO home has a highly structured psycho-social intervention program which includes group therapy, life skills training, occupational therapy, vocational training, horticulture and gardening, relaxation and games, social and cultural activities, which is far more than what an average individual with schizophrenia receives in the community as part of non-pharmacological treatment. Many patients in this category remain heavily dependent on the supportive residential facility. The majority are categorised as those with high dependency and poor capacity, and unable to live independently or earn. The persistence of severe disability in spite of rigorous supervised pharmacotherapy and nonpharmacological interventions again point to the unique clinical characteristics of this group.

The study also brings to focus the need for a clear plan of action for rehabilitation of these patients like in high-income countries ^(39).^ The new Mental Health Care Act 2017 in India puts the onus of providing adequate community based housing onto the government^(40).^ These are the patients who stay in government run hospitals (asylums of the bygone era) and who are incapable of living independently in the community. Studies have looked at various interventions for addressing homelessness in serious mentally ill population and its outcomes in different countries ^(41,42,43)^.Social housing projects fully funded by the government^(44,45)^ is the urgent need of the hour to ensure safe and secure supervised housing options for this population. In a country like India which aspires to rapidly progress in many social indices, supported housing for the serious mentally ill who are homeless need to be prioritised as the burden of care is huge in absolute numbers. This will go a long way in ensuring better outcomes in this most under-served patient population^(46,47)^, who tragically appears to have the most severe form of the illness.

### Limitations

Our study being observational, has its limitations in drawing conclusions regarding factors associated with homelessness. The sampling of patients was determined by their presence in the NGO and ability to provide informed consent. Additionally, the scores on symptom severity among women with schizophrenia living with their family were low, but comparable to that reported in other studies from the area^(48,49)^. The evaluations were conducted by raters with experience in conducting psychiatric assessments. The fact that the sample comprised exclusively of women, and were earlier in the course of illness may account for lower scores. It is also possible that women under-report symptoms because of stigma and social pressure to present as being well.

On the other hand, longitudinal studies, using imaging biomarkers in first episode psychosis, would help to confirm whether patients with homelessness represent a non-dopamine subtype of schizophrenia. Another limitation is that most of the homeless patients had no details regarding their age of onset of illness, duration of untreated psychosis (DUP) and could not provide an accurate treatment history. This limited making conclusions regarding non-response to psychotropic medications, even though supervised treatment for at least 6 months was ensured.

## Conclusions

The study shows that women with schizophrenia and homelessness represent an extreme phenotype of the illness, as reflected in greater symptom severity, cognitive deficits and disability. The study raises many questions which need to be answered through long term follow up studies. Is there a causal association between homelessness and disability? Is homelessness a cause or an effect of decline in functional ability? Are we looking at an extreme phenotype of hebephrenic schizophrenia with its unique treatment resistant features? Is there a non-dopamine mediated psychosis group? If homelessness as an outcome needs to be prevented, what would the most reliable intervention at the beginning of the illness in this group? Newer drugs that act on non-dopamine pathways, early identification, unique biomarker that predicts the type of illness and prognosis may be some of the most important requisites to reduce the burden of illness in this group of severely unwell patients. The major finding in the homeless population needs to be replicated in similar cohorts across the world with varying social and mental health support networks which would validate the hypothesis of an extreme phenotype of schizophrenia. Finally, the study also highlights the importance of incorporating ethnic and population diversity in schizophrenia research ^(50)^.

## Data Availability

All data produced in the present study are available upon reasonable request to the authors

## Disclosures

Rajiv Radhakrishnan is funded by VA National Center on Homelessness Among Veterans (RFQ36C24820Q1276), National Institute of Mental Health (NIMH) (R21MH123870) National Institute of Drug Abuse (NIDA), National Center for Complementary and Integrative Health (NCCIH) and has received research support from GW Pharmaceuticals (Jazz Pharmaceuticals) and Neurocrine Biosciences. The views expressed are those of the authors and not of those of the federal government or funding agencies.

Jayakumar Menon has worked as sub-investigator in a multi-national drug trial by Pfizer.

## References

1) Priebe S. Social outcomes in schizophrenia. The British Journal of Psychiatry. 2007;191(S50):s15–s20. doi:10.1192/bjp.191.50.s15

2) Davidson L, McGlashan TH. The Varied Outcomes of Schizophrenia. Can J Psychiatry. 1997;42(1):34–43. doi:10.1177/070674379704200105

3) Jauhar S, Johnstone M, McKenna PJ. Schizophrenia. Lancet. 2022;399(10323):473–486. doi:10.1016/S0140-6736(21)01730-X

4) Smartt C, Prince M, Frissa S, Eaton J, Fekadu A, Hanlon C. Homelessness and severe mental illness in low- and middle-income countries: scoping review. BJPsych Open. 2019;5(4):e57. doi:10.1192/bjo.2019.32

5) Tripathi A, Nischal A, Dalal PK, et al. Sociodemographic and clinical profile of homeless mentally ill inpatients in a north Indian medical university. Asian J Psychiatr. 2013;6(5):404–409. doi:10.1016/j.ajp.2013.05.002

6) Opler LA, White L, Caton CL, Dominguez B, Hirshfield S, Shrout PE. Gender differences in the relationship of homelessness to symptom severity, substance abuse, and neuroleptic noncompliance in schizophrenia. J Nerv Ment Dis. 2001;189(7):449–456. doi:10.1097/00005053-200107000-00006

7) Opler LA, Caton CL, Shrout P, Dominguez B, Kass FI. Symptom profiles and homelessness in schizophrenia. J Nerv Ment Dis. 1994;182(3):174–178. doi:10.1097/00005053-199403000-00008

8) Gupta S, Sagar R. National Mental Health Programme-Optimism and Caution: A Narrative Review. Indian Journal of Psychological Medicine. 2018;40(6):509–516. doi:10.4103/IJPSYM.IJPSYM_191_18

9) Ul Hassan F, Nagavarapu LS, Prasad M K, Raj A, Sekhar K. Homelessness in mental illness: Opportunities & prospects in the Indian context. Asian J Psychiatr. 2019;45:28–32. doi:10.1016/j.ajp.2019.08.011

10) Gowda GS, Telang A, Sharath CR, et al. Use of newer technologies with existing service for family reintegration of unknown psychiatric patients: A case series. Asian J Psychiatr. 2019;43:205–207. doi:10.1016/j.ajp.2017.10.022

11) Thara R, Kamath S. Women and schizophrenia. Indian J Psychiatry. 2015;57(Suppl 2):S246–S251. doi:10.4103/0019-5545.161487

12) Rao PN. Rehabilitation of the wandering seriously mentally ill (WSMI) women-The Banyan experience. Soc Work Health Care. 2004;39(1-2):49–65. doi:10.1300/j010v39n01_05

13) Andreasen NC. The Scale for the Assessment of Negative Symptoms (SANS): Conceptual and Theoretical Foundations. The British Journal of Psychiatry. 1989;155(S7):49–52. doi:10.1192/S0007125000291496

14) Nasreddine ZS, Phillips NA, Bédirian V, et al. The Montreal Cognitive Assessment, MoCA: a brief screening tool for mild cognitive impairment. J Am Geriatr Soc. 2005;53(4):695–699. doi:10.1111/j.1532-5415.2005.53221.x

15) Storey JE, Rowland JTJ, Basic D, Conforti DA, Dickson HG. The Rowland Universal Dementia Assessment Scale (RUDAS): a multicultural cognitive assessment scale. Int Psychogeriatr. 2004;16(1):13–31. doi:10.1017/s1041610204000043

16) Hurtado-Pomares M, Carmen Terol-Cantero M, Sánchez-Pérez A, Peral-Gómez P, Valera-Gran D, Navarrete-Muñoz EM. The frontal assessment battery in clinical practice: a systematic review. Int J Geriatr Psychiatry. 2018;33(2):237–251. doi:10.1002/gps.4751

17) Gil-Berrozpe GJ, Sánchez-Torres AM, García de Jalón E, et al. Utility of the MoCA for cognitive impairment screening in long-term psychosis patients. Schizophr Res. 2020;216:429–434. doi:10.1016/j.schres.2019.10.054.

18) Konecky B, Meyer EC, Marx BP, Kimbrel NA, Morissette SB. Using the WHODAS 2.0 to Assess Functional Disability Associated With DSM-5 Mental Disorders. AJP. 2014;171(8):818–820. doi:10.1176/appi.ajp.2014.14050587

19) Basavarajappa C, Kumar KS, Suresh VC, et al. What Score in WHODAS 2.0 12-Item Interviewer Version Corresponds to 40 % Psychiatric Disability? A Comparative Study Against IDEAS. J Psychosoc Rehabil Ment Health. 2016;3(1):21–26. doi:10.1007/s40737-016-0053-x

20) Reichenberg A, Velthorst E, Davidson M. Cognitive impairment and psychosis in schizophrenia: independent or linked conditions? World Psychiatry. 2019;18(2):162–163. doi:10.1002/wps.20644

21) Heinrichs RW, Zakzanis KK. Neurocognitive deficit in schizophrenia: A quantitative review of the evidence. Neuropsychology. 1998;12:426–445. doi:10.1037/0894-4105.12.3.426

22) Davidson L, McGlashan TH. The Varied Outcomes of Schizophrenia. Can J Psychiatry. 1997;42(1):34–43. doi:10.1177/070674379704200105

23) Llerena K, Gabrielian S, Green MF. Clinical and cognitive correlates of unsheltered status in homeless persons with psychotic disorders. Schizophr Res. 2018;197:421–427. doi:10.1016/j.schres.2018.02.023

24) Minzenberg MJ, Laird AR, Thelen S, Carter CS, Glahn DC. Meta-analysis of 41 Functional Neuroimaging Studies of Executive Function in Schizophrenia. Archives of General Psychiatry. 2009;66(8):811–822. doi:10.1001/archgenpsychiatry.2009.91

25) Weickert TW, Goldberg TE, Gold JM, Bigelow LB, Egan MF, Weinberger DR. Cognitive Impairments in Patients With Schizophrenia Displaying Preserved and Compromised Intellect. Archives of General Psychiatry. 2000;57(9):907–913. doi:10.1001/archpsyc.57.9.907

26) Reichenberg A (Avi). The assessment of neuropsychological functioning in schizophrenia. Dialogues in Clinical Neuroscience. 2010;12(3):383–392. doi:10.31887/DCNS.2010.12.3/areichenberg

27) Yamashita M, Shimokawa T, Takahashi S, et al. Cognitive functions relating to aberrant interactions between task-positive and task-negative networks: Resting fMRI study of patients with schizophrenia. Applied Neuropsychology: Adult. 2022;29(5):1122–1130. doi:10.1080/23279095.2020.1852565

28) Maggini C, Dalle Luche R. An overview on Hebephrenia, a diagnostic cornerstone in the neurodevelopmental model of Schizophrenia. Hist Psychiatry. 2022;33(1):34–46. doi:10.1177/0957154X211062534

29) Medeiros AB, Descalço N, Santos CF, Gomes R, Pereira MV. Hebephrenic schizophrenia as a variant of frontotemporal dementia – the true dementia praecox? European Psychiatry. 2021;64(S1):S165–S165. doi:10.1192/j.eurpsy.2021.440

30) Fenton WS, McGlashan TH. Natural History of Schizophrenia Subtypes: I. Longitudinal Study of Paranoid, Hebephrenic, and Undifferentiated Schizophrenia. Archives of General Psychiatry. 1991;48(11):969–977. doi:10.1001/archpsyc.1991.01810350009002

31) Tsai J, Szymkowiak D, Radhakrishnan R. Antipsychotic Medication Prescriptions for Homeless and Unstably Housed Veterans in the Veterans Affairs Health Care System. J Clin Psychiatry. 2020;82(1):20m13372. doi:10.4088/JCP.20m13372

32) Horga G. Neuromelanin-Sensitive MRI as a Candidate Biomarker for Psychiatric Disorders. Biological Psychiatry. 2022;91(9):S59–S60. doi:10.1016/j.biopsych.2022.02.168

33) Cassidy CM, Zucca FA, Girgis RR, et al. Neuromelanin-sensitive MRI as a noninvasive proxy measure of dopamine function in the human brain. Proc Natl Acad Sci U S A. 2019;116(11):5108–5117. doi:10.1073/pnas.1807983116

34) Wieland L, Fromm S, Hetzer S, Schlagenhauf F, Kaminski J. Neuromelanin-Sensitive Magnetic Resonance Imaging in Schizophrenia: A Meta-Analysis of Case-Control Studies. Front Psychiatry. 2021;12:770282. doi:10.3389/fpsyt.2021.770282

35) Giessen E van de, Pluijm M van der, Booij J, Haan L de. Neuromelanin MRI as Biomarker for Treatment Resistance in First Episode Schizophrenia. Biological Psychiatry. 2022;91(9):S61. doi:10.1016/j.biopsych.2022.02.171

36) Siskind D, McCartney L, Goldschlager R, Kisely S. Clozapine v. first- and second-generation antipsychotics in treatment-refractory schizophrenia: systematic review and meta-analysis. The British Journal of Psychiatry. 2016;209(5):385–392. doi:10.1192/bjp.bp.115.177261

37) Samara MT, Dold M, Gianatsi M, et al. Efficacy, Acceptability, and Tolerability of Antipsychotics in Treatment-Resistant Schizophrenia: A Network Meta-analysis. JAMA Psychiatry. 2016;73(3):199–210. doi:10.1001/jamapsychiatry.2015.2955

38) Sajatovic M, Ramirez LF, Fuentes-Casiano E, et al. A 6-month prospective trial of a personalized behavioral intervention + long-acting injectable antipsychotic in individuals with schizophrenia at risk for treatment non-adherence and homelessness. J Clin Psychopharmacol. 2017;37(6):702–707. doi:10.1097/JCP.0000000000000778

39) Montgomery AE, Metraux S, Culhane D. Rethinking Homelessness Prevention among Persons with Serious Mental Illness. Social Issues and Policy Review. 2013;7(1):58–82. doi:10.1111/j.1751-2409.2012.01043.x

40) https://egazette.nic.in/WriteReadData/2017/175248.pdf Mental Health Care Act 2017, Government of India

41) Gowda GS, Gopika G, Kumar CN, et al. Clinical outcome and rehabilitation of homeless mentally ill patients admitted in mental health institute of South India: “Know the Unknown” project. Asian J Psychiatr. 2017;30:49–53. doi:10.1016/j.ajp.2017.07.001

42) Kerman N, Sylvestre J, Aubry T, Distasio J, Schütz CG. Predictors of Mental Health Recovery in Homeless Adults with Mental Illness. Community Ment Health J. 2019;55(4):631–640. doi:10.1007/s10597-018-0356-3

43) Rosenheck R. Cost-effectiveness of services for mentally ill homeless people: the application of research to policy and practice. Am J Psychiatry. 2000;157(10):1563–1570. doi:10.1176/appi.ajp.157.10.1563

44) Folsom DP, Hawthorne W, Lindamer L, et al. Prevalence and Risk Factors for Homelessness and Utilization of Mental Health Services Among 10,340 Patients With Serious Mental Illness in a Large Public Mental Health System. AJP. 2005;162(2):370–376. doi:10.1176/appi.ajp.162.2.370

45) Stergiopoulos V, Mejia-Lancheros C, Nisenbaum R, et al. Long-term effects of rent supplements and mental health support services on housing and health outcomes of homeless adults with mental illness: extension study of the At Home/Chez Soi randomised controlled trial. The Lancet Psychiatry. 2019;6(11):915–925. doi:10.1016/S2215-0366(19)30371-2

46) Bighelli I, Rodolico A, García-Mieres H, et al. Psychosocial and psychological interventions for relapse prevention in schizophrenia: a systematic review and network meta-analysis. The Lancet Psychiatry. 2021;8(11):969–980. doi:10.1016/S2215-0366(21)00243-1

47) Smartt C, Prince M, Frissa S, Eaton J, Fekadu A, Hanlon C. Homelessness and severe mental illness in low- and middle-income countries: scoping review. BJPsych Open. 2019;5(4):e57. doi:10.1192/bjo.2019.32

48) Malla A, Iyer SN, Rangaswamy T, et al. Comparison of clinical outcomes following 2 years of treatment of first-episode psychosis in urban early intervention services in Canada and India. The British Journal of Psychiatry. 2020;217(3):514–520. doi:10.1192/bjp.2020.126

49) Ann-Catherine Lemonde and others, Differential Trajectories of Delusional Content and Severity Over 2 Years of Early Intervention for Psychosis: Comparison Between Chennai, India, and Montréal, Canada, Schizophrenia Bulletin, 2023;, sbad007, https://doi.org/10.1093/schbul/sbad007

50) Burkhard C, Cicek S, Barzilay R, Radhakrishnan R, Guloksuz S. Need for Ethnic and Population Diversity in Psychosis Research. Schizophr Bull. 2021;47(4):889–895. doi:10.1093/schbul/sbab048

